# Performance of Immunoglobulin G Serology on Finger Prick Capillary Dried Blood Spot Samples to Measure SARS-CoV-2 Humoral Immunogenicity

**DOI:** 10.1101/2021.07.29.21261156

**Authors:** Aidan M. Nikiforuk, Brynn McMillan, Sofia R. Bartlett, Ana Citlali Márquez, Tamara Pidduck, Jesse Kustra, David M. Goldfarb, Vilte Barakauskas, Graham Sinclair, David M. Patrick, Manish Sadarangani, Gina S. Ogilvie, Muhammad Morshed, Inna Sekirov, Agatha N. Jassem

## Abstract

**Importance:** Measuring humoral immunogenicity of Severe Acute Respiratory Syndrome Coronavirus 2 vaccines and finding population-level correlates of protection against coronavirus disease presents an immediate challenge to public health practitioners.

**Objective:** To study the diagnostic accuracy and predictive value of finger prick capillary dried blood spot samples tested using an anti-immunoglobulin G (IgG) serology assay to measure SARS-CoV-2 seropositivity and the humoral immunogenicity of COVID-19 vaccination.

**Design, Setting and Participants:** This cross-sectional study enrolled participants (n= 644) who had paired DBS and serum samples collected by finger prick and venipuncture, respectively, in British Columbia, Canada between January 12^th^, 2020 and May 21^st^, 2021. Samples were tested by a multiplex electrochemiluminescence assay for SARS-CoV-2 anti-Spike (S), -Nucleocapsid (N) and -receptor binding domain (RBD) IgG reactivity using a Meso Scale Discovery (MSD) platform. Additionally, unpaired DBS samples (n= 6,706) that were collected in the province during the same time period were included for analysis of SARS-CoV-2 anti-N IgG reactivity.

**Exposure:** Collection of a capillary dried blood spot by finger prick alone or paired with serum by venipuncture.

**Outcome:** Humoral immune response to SARS-CoV-2 measured by detection of anti-S, -N or - RBD IgG.

**Results:** In comparison to a paired-serum reference, dried blood spot samples possess a sensitivity of 80% (95% CI: 61%-91%) and specificity of 97% (95% CI: 95%-98%). Receiver operator characteristic curve analysis (ROC) found that participant DBS samples tested for anti-SARS-CoV-2 IgG by MSD V-PLEX COVID-19 Coronavirus Panel 2 assay accurately classify SARS-CoV-2 seroconversion at an 88% percent rate, AUC= 88% (95% CI: 81%-96%). Modelling found that a dried blood spot-based testing approach has a high positive predictive value (98% [95% CI: 98%-99%]) in a theoretical population with seventy-five percent COVID-19 vaccine coverage. At lower vaccine coverages of fifteen and forty-five percent, the test’s positive predictive value decreased, and the negative predictive value increased.

**Conclusion:** We demonstrate that dried blood spot collected samples, when tested using an electrochemiluminescence assay, provide a valid alternative to traditional venipuncture and should be considered to reliably detect SARS-CoV-2 seropositivity.

**Key Points:** *Question:* What is the diagnostic accuracy and predictive value of immunoglobulin G serology on finger prick capillary dried blood spot samples to measure SARS-CoV-2 humoral immunogenicity?

*Findings:* In comparison to a paired-serum reference, dried blood spot samples tested for anti-SARS-CoV-2 IgG possess a sensitivity of 80% (95% CI: 61%-91%) and specificity of 97% (95% CI: 95%-98%). Dried blood spot testing has a positive predictive value of 98% (95% CI: 98%-99%) when modelled in a theoretical population with COVID-19 vaccine coverage of seventy-five percent.

*Meaning:* Dried blood spot samples have equal diagnostic accuracy to serum collected by venipuncture when tested by electrochemiluminescence assay and should be considered to reliably detect SARS-CoV-2 seropositivity.

## Introduction

The ongoing global vaccination campaign to immunize populations against Severe Acute Respiratory Syndrome Coronavirus 2 (SARS-CoV-2) infection, which causes coronavirus disease (COVID-19), represents the largest primary prevention effort undertaken in public health since the Global Polio Eradication Initiative (GPEI)^1^. Lessons learned from the GPEI highlight the value of measuring population level vaccine elicited immunogenicity, as the humoral response can differ between doses, age groups, vaccine formulations and viral strains^1^.

The size and scale of the COVID-19 vaccination campaign raises logistical challenges in measuring the humoral immunogenicity of SARS-CoV-2 vaccination at the population level. The requirement for high volume serologic testing to detect vaccine elicited immunogenicity represents a particular challenge, as whole blood specimen collection by venipuncture does not easily scale up^2,3^. Whole blood collection in the form of serum or plasma requires a trained phlebotomist, specific collection tubes, and cold chain logistics. Dried blood spot (DBS) sampling is a cost effective and promising alternative, which can occur by self-collection, eliminating the need for trained personnel. Collection cards are stable at ambient temperatures for up to two weeks, simplifying transport, and can be stored in large quantities using less space^4^. To understand the potential of DBS samples in measuring SARS-CoV-2 seropositivity at the population level we: i) evaluated the diagnostic accuracy of DBS tested by Meso Scale Discovery (DBS-MSD) multiplex anti-IgG electrochemiluminescence assay in comparison to a paired-serum reference in study participants (n= 644; 30 positive and 614 negative) from British Columbia, Canada and, ii) modelled the predictive performance of DBS-MSD testing in a theoretical population (n=10,000) with stratified COVID-19 vaccine coverage of fifteen, forty-five and seventy-five percent.

## Methods

Paired participant samples (n= 644) were received at the British Columbia Centre for Disease Control Public Health Laboratory. Serum samples were collected from venipuncture by trained phlebotomists in 5ml tubes (BD Vacutainer^®^ SST(tm) Tubes: #367986), centrifuged and tested before storage at −20°C. DBS samples were collected by capillary finger-prick using a contact activated lancet (BD Microtainer^®^: #366594), spotted on protein saver cards (Whatman 903^(tm)^: #Z761575), sealed in a gas-impermeable sachet with 1gm of desiccant per card, and stored at −20°C. DBS sample collection was performed by a health care worker or by self-collection. Written instructions were provided to participants who were asked to self-collect. Four 6mm punches were eluted in dipotassium phosphate buffered saline with 0.5% sodium azide and 1.5% bovine serum albumin w/v (Ortho Clinical Diagnostics, personal communication). Serum samples were diluted 1:5000 v/v and DBS samples were diluted 1:500 v/v in Diluent 100 (MSD: #R50AA-2) before serological testing. Serological testing was performed with the V-PLEX COVID-19 Coronavirus Panel 2 (IgG) (MSD: #K15369U), offered by MSD^5^. Reference positive samples were defined as the paired serum samples that had signals above the MSD recommended target thresholds for anti-Spike (S), and/or -Nucleocapsid (N) and/or -receptor binding domain (RBD) IgG (S=1960, N=5000 and RBD=538)^5^. Serum samples were classified as positive for SARS-CoV-2 anti-IgG when signal for two of three targets was greater-than or equal-to the threshold. Thresholds were set for DBS-MSD results by plotting distributions of anti-S and -N IgG signals. DBS-MSD samples were classified as positive if the anti-S and -N signal or only the anti-S signal was greater-than or equal-to the thresholds established for DBS samples. Nucleocapsid-only positive DBS-MSD samples were classified as negative. Anti-RBD results were not interpreted for DBS samples, as they were found to be highly correlated with anti-S signals (Pearson’s correlation, r = 95%, 95% CI [92%-97%]), indicating collinearity^6^. Sensitivity and specificity of DBS-MSD was calculated in comparison to the paired-serum reference. Logistic regression was used to predict the paired-serum reference diagnosis by DBS-MSD result^7^. Internal cross-validation was performed by a n= 2,000 bootstrap and results were plotted as a receiver operator characteristic curve (ROC)^8,9^. The positive- and negative-predictive value (PPV, NPV) of DBS-MSD was modelled in a theoretical population of ten-thousand persons (n= 10,000) with varying COVID-19 vaccine coverage of fifteen, forty-five and seventy-five percent^10,11^.

The University of British Columbia Clinical Research Ethics Board provided ethical review and approval for studies from which participants’ specimens were cross-sectionally sampled and tested (H20-02184, H20-02402, H20-01421 and H20-01886).

## Results

Signal target thresholds for DBS-MSD were established at 75 AU/ml (95% CI: 55-95 AU/ml) for anti-S (Figure 1a) and 175 AU/ml (95% CI: 162-188 AU/ml) anti-N IgG (Figure 1b), based on the observed distribution of signals in DBS samples. The anti-S and anti-N thresholds were set to maximize sensitivity and specificity, respectively. The anti-S distribution shows that 77% of SARS-CoV-2 serum positive samples have a DBS signal greater than or equal to the threshold (one sample T-test, P=0.77, Sensitivity= 77%). The anti-N distribution estimates that 95% of anti-S negative DBS samples have a signal less than or equal to the threshold (one sample T-test, P=0.05, Specificity= 95%). Applying these thresholds to the same dataset (n= 644), DBS-MSD achieved a sensitivity of 80% (95% CI: 61%-91%) and specificity of 97% (95% CI: 95%-98%) in comparison to the paired-serum reference (Figure 2a). Internally cross-validated ROC analysis yielded an area under the curve of 88% (95% CI: 81%-96%) (Figure 2b). DBS samples resulted on MSD will accurately classify SARS-CoV-2 seroconversion at an 88% percent rate. In a theoretical population of 10,000 persons, of which seventy-five percent have received COVID-19 immunization, a positive test result predicts true seropositivity at a 98% rate, PPV= 98% (95% CI: 98%-99%) (Figure 3). A negative test result predicts a seronegative response at a 38% rate, NPV= 38% (95% CI: 37%-40%). At lower vaccine coverages of fifteen and forty-five percent, the PPV of DBS-MSD decreased and the NPV increased (PPV_15_ = 29% (95% CI: 25%-33%), NPV_15_ = 99% (95% CI: 99%-100%) and PPV_45_ = 96% (95% CI: 95%-96%), NPV_45_ = 86% (95% CI: 85%-86%).

**Figure 1:**
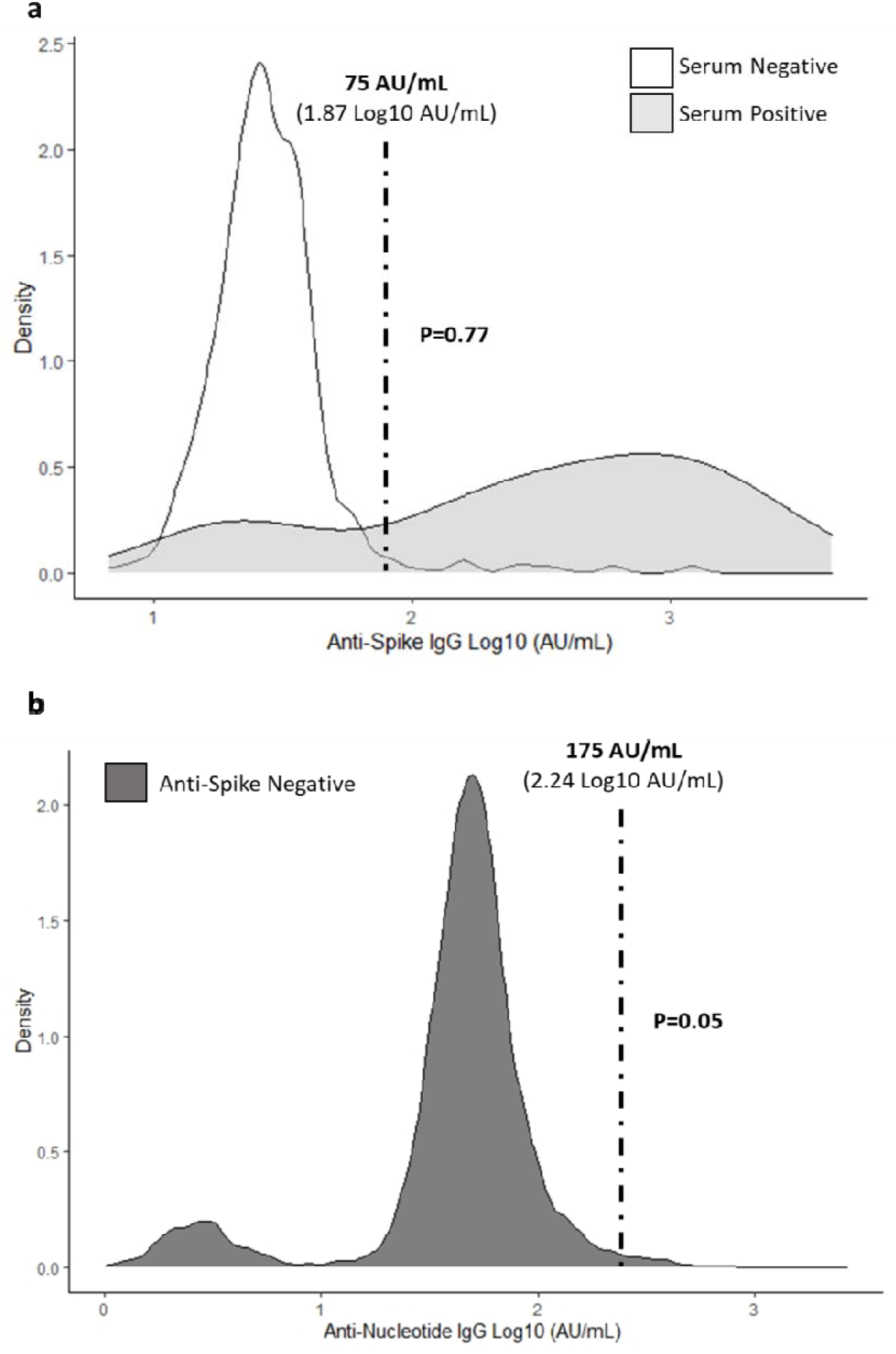
Signal distributions of SARS-CoV-2 anti-Spike (S) and -Nucleocapsid (N) IgG collected by DBS and tested with an MSD assay. **a)** Participant DBS and paired-serum samples n=644, were tested by MSD assay for anti-S, -N and -receptor binding domain (RBD) IgG. DBS-MSD anti-S signals were stratified by MSD test-results on paired-serum samples (white: paired-serum positive) (grey: paired-serum negative). A sample was classified as paired-serum positive when greater-than or equal-to two of three target signals exceeded the manufacturer recommended thresholds (S= 1960, N= 5000 and RBD= 538). A threshold of ≥ 75 AU/ml (95% CI: 55-95 AU/ml) was set for anti-S DBS samples tested on MSD, as it discriminates paired-serum positives from negatives. In a random sample of serum positives, 77% of paired-DBS-MSD samples are expected to have values greater than or equal to 75 AU/ml (one sample T-test, P= 0.77, Sensitivity= 77%). **b)** All anti-N DBS-MSD samples tested at the BCCDC to May 21^st^, 2021 were restricted to those with DBS-MSD anti-S < 75 AU/ml (n= 6,706) (dark grey). A threshold of ≥ 175 AU/ml (95% CI: 162-188 AU/ml) was set for anti-N DBS samples tested on MSD, as the probability of classifying an anti-S negative DBS-MSD sample anti-N positive equals 5% (one sample T-test, P= 0.05, Specificity= 95%). DBS-MSD samples were classified positive if anti-S signal was ≥ 75 AU/ml and anti-N signal was ≥ 175 AU/ml or anti-S signal was ≥ 75 AU/ml and anti-N signal was < 175 AU/ml. Samples with anti-S signal < 75 AU/ml and anti-N signal ≥ 175 AU/ml were classified as negative.

**Figure 2:**
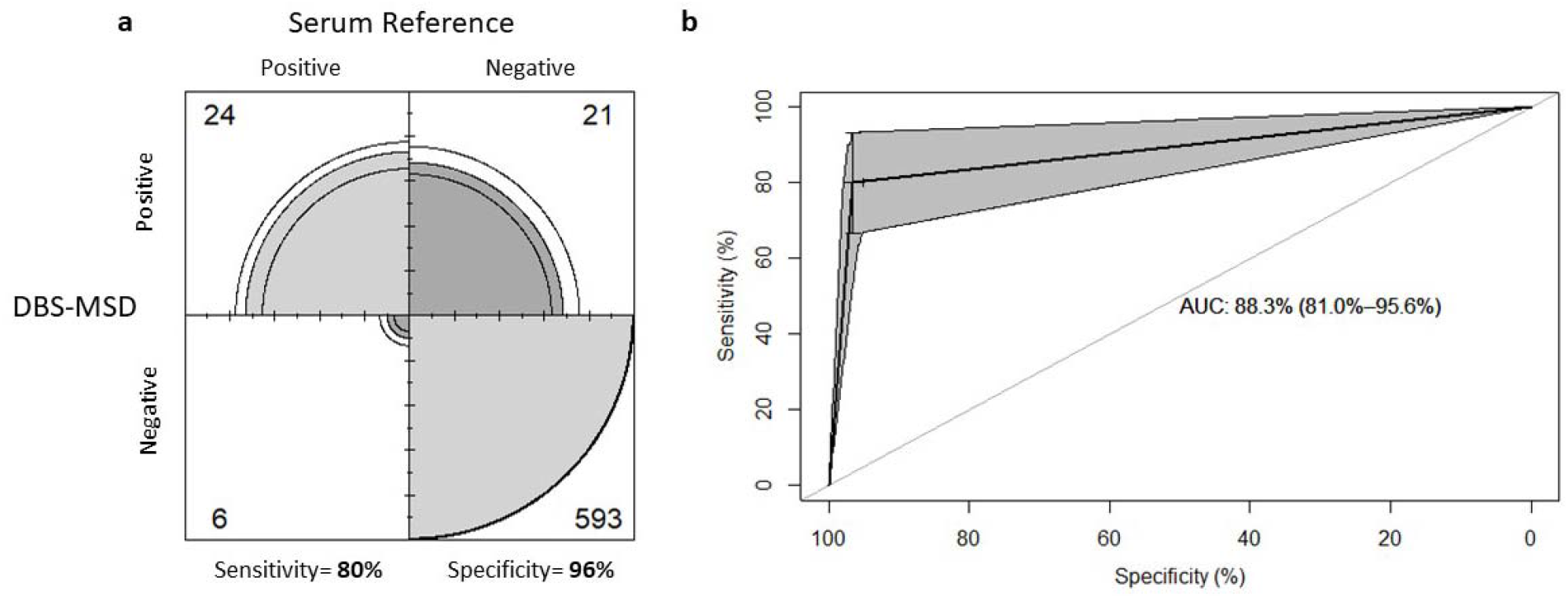
Confusion matrix and internally cross-validated receiver operating characteristic curve analysis of DBS-MSD test result in comparison to the paired-serum reference. **a)** Frequency of DBS-MSD results are reported in comparison to the reference and used to calculate clinical accuracy (sensitivity and specificity). In comparison to the paired-serum reference, DBS-MSD possesses a sensitivity of 80% (95% CI: 61%-91%) and specificity of 97% (95% CI: 95%-98%), grey area shows the proportion of participants by cell and black lines represent the 95% confidence interval. No evidence of similarity between the marginal outcome probability was observed (McNemar test, P<0.007). **b)** Internally cross-validated receiver operator characteristic curve analysis was used to quantify the predictive ability of a DBS-MSD test in comparison to the reference. DBS-MSD was found to accurately discriminate natural SARS-CoV-2 seroconversion at an 88% (95%CI: 81%-96%) rate (bootstrap= 2000).

**Figure 3:**
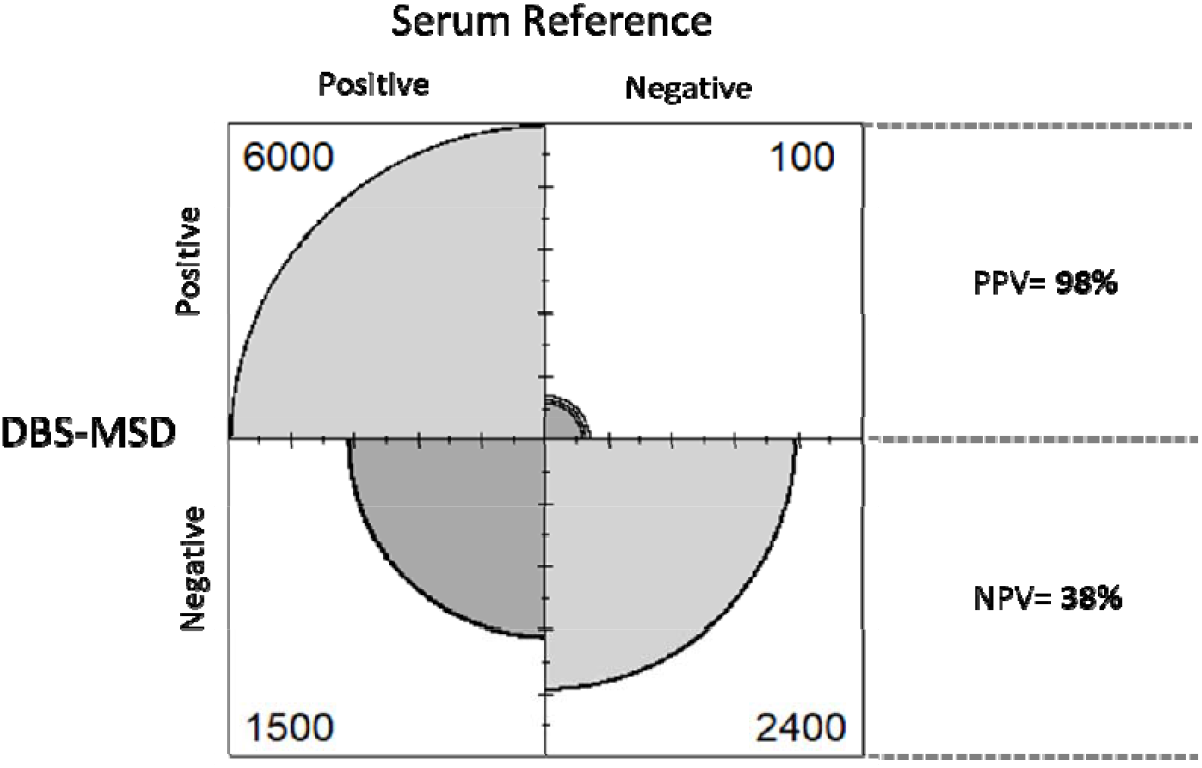
Confusion matrix showing the predictive performance of MSD-DBS in a theoretical population (n=10,000) with seventy-five percent SARS-CoV-2 vaccine coverage. The positive-predictive value (PPV) of MSD-DBS is positively correlated with the prevalence of SARS-CoV-2 seroreactivity. At seventy-five percent SARS-CoV-2 vaccine coverage, a positive MSD-DBS test would correctly identify vaccine elicited seroreactivity at a 98% rate, PPV= 98% (95% CI: 98%-99%). A negative MSD-DBS test correctly identifies no vaccine elicited seroreactivity at a 38% rate, negative predictive value (NPV)= 38% (95% CI: 37%-40%). There was no evidence of independence between results (Chi^2^ test, P<0.001).

## Discussion

The diagnostic accuracy of DBS-MSD is comparable to that of the Roche Elecsys^®^ platform for surveillance of SARS-CoV-2 seroconversion^12^. Tests performed on DBS samples characteristically exhibit high specificity and low sensitivity, attributable to low analyte concentration, variance in sample collection practices or time since antigen exposure^13^. We show that, despite the expected disadvantage of low analyte concentration, DBS-MSD testing may possess a strong PPV when implemented in a context with high SARS-CoV-2 seroprevalence (e.g., post a successful COVID-19 immunization campaign). Therefore, DBS collection and anti-IgG serology shows promise as a tool to measure SARS-CoV-2 seropositivity at the population level. The benefits include comparable diagnostic accuracy to that of serum samples, with improved population reach, and the potential to discriminate natural from vaccine elicited seroconversion by measuring anti-S and anti-N IgG reactivity^14^. In a theoretical population with seventy-five percent COVID-19 vaccine coverage, a positive DBS-MSD result reliably indicates seropositivity with a high PPV and does not require additional testing. The low NPV indicates that a negative result does not reliably predict lack of an immune response or immunosenescence and may require reflex testing. In receipt of a negative result, where confirmation of individual serostatus is required, reflex testing may entail additional testing from venipuncture or DBS.

An important limitation of SARS-CoV-2 seroprevalence studies in populations with low vaccine coverage is the difference in antibody signals between individuals who have recovered from asymptomatic and symptomatic infection^15^. The distribution of serological signals between recovered asymptomatic and negative cases are more likely to overlap than when comparing negative to recovered symptomatic, nucleic acid amplification test confirmed cases. Low serological signal should not be a limitation in populations with high vaccine coverage, as SARS-CoV-2 vaccination has been experimentally found to elicit a stronger humoral immune response than natural infection^16^.

In summary, measuring SARS-CoV-2 seropositivity at the population level presents unique challenges, which warrant investigation and consideration of alternative methodologies. We show robust diagnostic accuracy of DBS samples when tested for anti-SARS-CoV-2 IgG using an MSD assay, and model the predictive value of DBS-MSD testing in a theoretical population with fifteen, forty-five and seventy-five percent COVID-19 vaccine coverage^11^. DBS tests have comparable specificity and lower sensitivity to those conducted on serum, due to low analyte volume and the need for additional sample processing. The PPV of DBS-MSD testing increases in response to high seroprevalence, making it possible to accurately identify individuals who have a humoral immune response. Follow-up, reflex testing may be required to confirm true negatives. The sensitivity and NPV of DBS-MSD testing will likely increase in an immunized population, as vaccinated individuals have a more robust humoral response than those that are naturally infected^16^.

At the population level, naturally infected and vaccinated individuals can be considered a homogenized group, where an anti-S IgG positive signal indicates seroreactivity. Conversely, individual diagnosis may require further interpretation where anti-S IgG positive persons are further stratified by their anti-N IgG results and/or clinical information (e.g., vaccination status, prior laboratory results) to determine natural infection from vaccine elicited immunity. Detection of natural infection by serology alone is dependent on study design, as anti-N IgG signal may wane when the time between exposure and sample collection lengthens.

Public health practitioners should consider the utility of DBS-MSD to evaluate SARS-CoV-2 seropositivity from natural infection or COVID-19 vaccination. Modelling shows this combination possesses a strong PPV (∼ 98%) in settings of high seroprevalence (e.g., seventy-five percent COVID-19 vaccine coverage). Public health agencies are challenged with simultaneously administering COVID-19 vaccines and measuring the elicited immune response. To address the later, a reliable and accessible method for detecting SARS-CoV-2 seropositivity is required. The logistic, economic and demonstrated diagnostic accuracy of DBS-MSD make it a strong candidate for population level investigation of SARS-CoV-2 humoral immunogenicity, especially in longitudinal study designs requiring repeated measures.

## Data Availability

The data that support the findings of this study are available from the data steward but restrictions apply to the availability of these data, which were used under license for the current study, and so are not publicly available. Data are however available from the authors upon reasonable request and with permission of the data steward.

## Acknowledgements

This study was funded by the Michael Smith Foundation for Health Research (COV-2020-1120, COV-2020-1279) and Genome British Columbia (COV-050). Additional funding was provided by the Public Health Agency of Canada through the COVID-19 Immunity Taskforce (2021-HQ-000141), BC SUPPORT Unit (C19-PE-V4), Canadian Institutes of Health Research (#434951) and the University of British Columbia-Public Scholars Initiative. M.S. is supported via salary awards from the BC Children’s Hospital Foundation, the Canadian Child Health Clinician Scientist Program and the Michael Smith Foundation for Health Research.

## Contributions

A.M.N. contributed to attaining funding, performed serological testing, data analysis, wrote and edited the manuscript.

B.M. assisted with sample acquisition, planning, performed serological testing, acquiring and interpreting data. B.M. assisted in writing and editing the manuscript.

S.R.B. contributed to attaining funding, designing the study, sample collection, serological testing. S.B. discussed the results of the data analysis and assisted in interpreting the results.

A.C.M. assisted in planning experiments, data analysis and interpretation of the results. T.P. and J.K. assisted with sample acquisition, planning and performing experiments, and acquiring data.

G.D. and V.B. assisted in collecting paired samples from participants at the CW campus and evaluation of validation data.

G.S. and V.B. assisted with study design related to DBS collection and testing.

D.M.P. assisted in data interpretation and edited the manuscript.

M.S., G.S.O., M.M., I.S. contributed to the study design, data interpretation and edited the manuscript.

A.N.J. obtained grant funding, contributed to the study design and assisted in interpreting the results, writing and editing the manuscript.

## Conflict of Interest Disclosures

S.R.B. has advised and spoken for Gilead Sciences and AbbVie (all personal payments given as unrestricted donations to BC Centre for Disease Control Foundation for Public Health) and has received investigator-initiated research funding from Gilead Sciences via her institution. M.S. has been an investigator on projects funded by GlaxoSmithKline, Merck, Pfizer, Sanofi-Pasteur, Seqirus, Symvivo and VBI Vaccines. All funds have been paid to his institute, and he has not received any personal payments.

## Notes

### Author Declarations

The University of British Columbia Clinical Research Ethics Board provided ethical review and approval for studies from which participants' specimens were cross-sectionally sampled and tested (H20-02184, H20-02402 and H20-01886).

